# Molecular Epidemiology of SARS-CoV-2 in Cyprus

**DOI:** 10.1101/2021.03.16.21252974

**Authors:** Jan Richter, Pavlos Fanis, Christina Tryfonos, Dana Koptides, George Krashias, Stavros Bashiardes, Andreas Hadjisavvas, Maria Loizidou, Anastasis Oulas, Denise Alexandrou, Olga Kalakouta, Mihalis I. Panayiotidis, George M. Spyrou, Christina Christodoulou

## Abstract

Whole genome sequencing of viral specimens following molecular diagnosis is a powerful analytical tool of molecular epidemiology that can critically assist in resolving chains of transmission, identifying of new variants or assessing pathogen evolution and allows a real-time view into the dynamics of a pandemic. In Cyprus, the first two cases of COVID-19 were identified on March 9, 2020 and since then 33,567 confirmed cases and 230 deaths were documented. In this study, viral whole genome sequencing was performed on 133 SARS-CoV-2 positive samples collected between March 2020 and January 2021. Phylogenetic analysis was conducted to evaluate the genomic diversity of circulating SARS-CoV-2 lineages in Cyprus. 15 different lineages were identified that clustered into three groups associated with the spring, summer and autumn/winter wave of SARS-CoV2 incidence in Cyprus, respectively. The majority of the Cypriot samples belonged to the B.1.258 lineage first detected in September that spread rapidly and largely dominated the autumn/winter wave with a peak prevalence of 86% during the months of November and December. The B.1.1.7 UK variant (VOC-202012/01) was identified for the first time at the end of December and spread rapidly reaching 37% prevalence within one month. Overall, we describe the changing pattern of circulating SARS-CoV-2 lineages in Cyprus since the beginning of the pandemic until the end of January 2021. These findings highlight the role of importation of new variants through travel towards the emergence of successive waves of incidence in Cyprus and demonstrate the importance of genomic surveillance in determining viral genetic diversity and the timely identification of new variants for guiding public health intervention measures.

## Introduction

Corona Virus Disease-2019 (COVID-19) is a pulmonary disease caused by the Severe Acute Respiratory Syndrome Coronavirus 2 (SARS-CoV-2) coronavirus [1]. SARS-CoV-2 was first detected in December 2019 in Wuhan city in the Hubei province of China in a patient with acute pneumonia [2,3]. Following global spread of the virus, on March 11, 2020 the World Health Organization (WHO) characterized COVID-19 as a pandemic, which is still ongoing and as of February 24, 2021 over 110 million confirmed cases and 2.47 million deaths were reported worldwide (https://covid19.who.int/).

The complete genome of SARS-CoV-2 isolated from a patient in Wuhan, China was initial published on Jan 5, 2020 [2] and since then analysis of viral sequences worldwide is continuous with more than 614,000 complete genomes currently available in public databases such as the GISAID (https://www.gisaid.org/). The global real-time tracing of the viral spread through whole genome sequencing is important in the race to timely identify the emergence of novel SARS-CoV-2 variants that change the transmission, antigenic properties and/or pathogenicity of the virus [4]. The recent identification of three major Variants of Concern (VOC), with increased transmissibility and the potential to reduce vaccine effectiveness has led to increased surveillance efforts worldwide [5,6].

The Republic of Cyprus is one of the countries in Europe least affected by the COVID-19 pandemic, presumably due to its rapid and effective response strategy that included high number of COVID-19 tests, effective tracing and isolation of cases and their contacts together with preventive measures (e.g. social distancing, wearing face masks and hand washing) [7]. In addition, being an island guaranteed high effectiveness of airport closure with regard to importing of new cases. The first two cases of COVID-19 in Cyprus were identified on March 9, 2020 and since then 33,567 confirmed cases and 230 deaths were documented (https://covid19.ucy.ac.cy/).

The aim of our study was to analyse and evaluate the genomic diversity of circulating SARS-CoV-2 lineages in Cyprus. We report 133 full genome sequences obtained from individuals tested positive by RT-PCR between March 2020 and January 2021 and evaluate their characteristics and relationship with the progression of the pandemic in Cyprus. Public sharing and use of genomic analyses are important tools for disease surveillance systems to track and trace acquired COVID-19 cases for more accurate decision making and appropriate public health action.

## Materials and Methods

### Sample collection

The Department of Molecular Virology of Cyprus Institute of Neurology and Genetics was assigned as the reference laboratory for SARS-CoV2 by the Cyprus Ministry of Health of the Republic of Cyprus. More than 180,000 samples from public as well as private hospitals were received and analysed since March 2020. For this retrospective observational study 133 SARS-CoV2 positive nasopharyngeal swabs obtained from patients referred for diagnostic purposes between March 2020 and January 2021 were selected for full genome sequencing. A random, unselected approach for sample selection was taken without pre-screening for variants of interest to avoid sampling bias. The viral RNA had been previously detected using a qRT-PCR assay and all samples selected had a cycle threshold value (Ct) lower than 30. The study was approved by the Cyprus National Bioethics committee (EEBK 21.1.01.03).

### Next Generation Sequencing

Total RNA was extracted from 200 μl of nasopharyngeal swab fluid in a final volume of 50 μl, using the MagMAX Viral/Pathogen Nucleic Acid Isolation Kit (Applied Biosystems) on a KingFisher™ Flex Purification System (Thermo Fisher Scientific). Libraries were prepared using the QIAseq SARS-CoV-2 Primer Panel in conjunction with the QIAseq FX DNA Library Kit (Qiagen) according to manufacturer’s instructions. In brief, viral RNA was reverse transcribed to synthesize cDNA using random hexamers. cDNAs were amplified in two high-fidelity multiplex PCR reactions using two different primer pools that together cover the entire SARS-CoV2 genome. The two enriched pools per sample were combined, purified using AMPure XP beads (Beckman Coulter) and quantified using the Qubit dsDNA HS Assay kit (Invitrogen). Fragmentation, end-repair and A-tailing was performed in a combined reaction per sample using 200 ng DNA. Next, Illumina platform-specific adapters were ligated to both ends of the DNA fragments. Library size selection and purification were carried out using AMPure XP beads (Beckman Coulter) in two rounds (0.8x and 1x respectively). The libraries were quality analyzed using the Agilent High Sensitivity D1000 ScreenTape on a 2200 TapeStation system (Agilent) and quantified using the Qubit dsDNA HS Assay kit (Invitrogen). Equimolar quantities of libraries were pooled (24 samples/pool) and sequenced on a Illumina MiSeq sequencer. All sequences obtained were deposited at the GISAID EpiCov database (www.gisaid.org).

### Mapping, alignment and lineage assignment

The Burrows-Wheeler Aligner (BWA), version: 0.7.15 was used to map the raw reads to the coronavirus reference genome Wuhan-Hu-1 (NCBI ID:NC_045512.2) [8]. Duplicate reads, which are likely to be the results of PCR bias, were marked using Picard version: 2.6.0 (http://broadinstitute.github.io/picard/). Samtools, version: 0.1.19, was used for additional BAM/SAM file manipulations [9]. SNPs, amino acid replacements and indels were verified using the CoV-Glue web interface [10]. Aligned reads were validated using the Integrative Genomics Viewers v.2.9.2 and consensus sequences were extracted [11]. SARS-CoV2 lineages of the 133 sequences were assigned using the dynamic nomenclature tool PANGOLIN (https://pangolin.cog-uk.io) [12].

### Phylogenetic analysis

All phylogenetic analyses were conducted in MEGA7 [13]. The alignment of consensus sequences was performed using MUSCLE. Bayesian information criterion (BIC) scores were calculated for different models to determine the best fitting nucleotide substitution model. In addition, jModelTest [14] was used for evaluating the best fitting nucleotide substitution model under BIC yielding the same result (TN93model + gamma distributed rates). This model was subsequently used to construct a Maximum Likelihood phylogenetic tree with 1000 bootstrap replicates. All positions containing alignment gaps and missing data were eliminated only in pairwise sequence comparisons.

## Results and Discussion

Since the beginning of the pandemic and until the 24/2/2021, the Republic of Cyprus reported 33,567 SARS-CoV2 confirmed cases (3.63% of the population) along with 230 associated casualties (24.2/100,000) (https://covid19.ucy.ac.cy/). Based on the number of positives identified (see Fig 1) the course of the pandemic in Cyprus so far can be distinguished into three main phases: a first phase during Spring 2020, starting with the first two cases identified on the 9^th^ of March that peaked in the first week of April that was contained with a nationwide lock-down and lasted approximately until June; a second phase during the summer that started after the re-opening of the airports; and a third phase (which is still continuing) during autumn/winter that peaked at the beginning of January and accounted for the majority of all cases registered so far.

**Fig 1.**
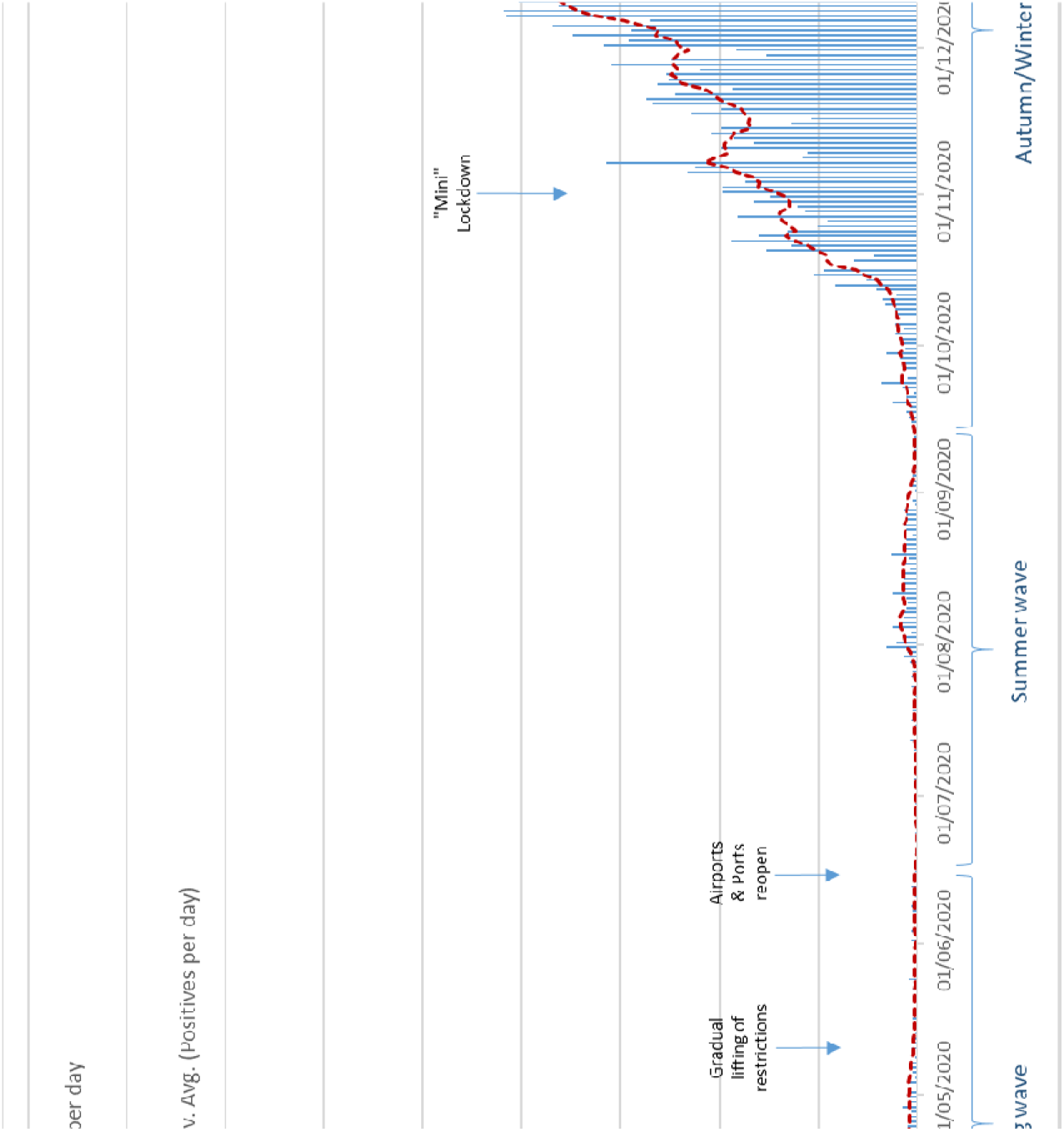
Daily number of positive SARS-CoV2 cases in Cyprus identified between March 2020 and January 2021 (https://covid19.ucy.ac.cy/). The start of the implementation as well as the lifting of important governmental pandemic control measures are indicated.

Overall, between the 11^th^ of March 2020 and the 29^th^ of January 2021, 15 different lineages were identified among the 133 samples sequenced (Table 1). As shown in Fig 3 the phylogenetic analysis is in support of the PANGOLIN lineages assignment and reveals a picture of independent distinct, importations that are followed by local transmission (Fig 3). In the bubble chart in Fig 2 three groups of lineages are clearly distinguishable and associate with the three phases of the SARS-CoV2 pandemic.

**Table 1.** Time course of SARS-CoV2 lineages identified in Cyprus between March 2020 and January 2021.

|  | Lineage | First detected | Last detected | Mar-20 | Apr-20 | May-20 | Jun-20 | Jul-20 | Aug-20 | Sep-20 | Oct-20 | Nov-20 | Dec-20 | Jan-21 | Total |
| --- | --- | --- | --- | --- | --- | --- | --- | --- | --- | --- | --- | --- | --- | --- | --- |
| Spring Wave | B.1.1.74 | 11/03/20 | 10/06/20 | 1 | 2 | 1 | 1 |  |  |  |  |  |  |  | 5 |
|  | B.1.1.277 | 22/03/20 | 25/03/20 | 3 |  |  |  |  |  |  |  |  |  |  | 3 |
|  | B.1.235 | 17/04/20 | 17/04/20 |  | 1 |  |  |  |  |  |  |  |  |  | 1 |
|  | B.1.1.251 | 27/04/20 | 05/05/20 |  | 1 | 1 |  |  |  |  |  |  |  |  | 2 |
| Summer wave | B.1.36 | 11/07/20 | 11/07/20 |  |  |  |  | 1 |  |  |  |  |  |  | 1 |
|  | B.1.236 | 17/07/20 | 18/08/20 |  |  |  |  | 5 | 2 |  |  |  |  |  | 7 |
|  | B.1.2 | 05/08/20 | 23/09/20 |  |  |  |  |  | 10 | 7 |  |  |  |  | 17 |
|  | B.1.1.67 | 13/08/20 | 13/08/20 |  |  |  |  |  | 1 |  |  |  |  |  | 1 |
|  | B.1.1.288 | 19/08/20 | 19/08/20 |  |  |  |  |  | 1 |  |  |  |  |  | 1 |
|  | B.1.1.192 | 27/08/20 | 27/08/20 |  |  |  |  |  | 1 |  |  |  |  |  | 1 |
| Autumn/Winter wave | B.1.177 | 16/09/20 | 26/01/21 |  |  |  |  |  |  | 1 | 3 | 2 | 1 | 2 | 9 |
|  | B.1.258 | 23/09/20 | 27/01/21 |  |  |  |  |  |  | 7 | 16 | 12 | 25 | 13 | 73 |
|  | B.1.1.103 | 28/12/20 | 29/12/20 |  |  |  |  |  |  |  |  |  | 2 |  | 2 |
|  | B.1.1.7 | 28/12/20 | 27/01/21 |  |  |  |  |  |  |  |  |  | 1 | 9 | 10 |
|  |  |  |  | 4 | 4 | 2 | 1 | 6 | 15 | 15 | 19 | 14 | 29 | 24 | 133 |

**Fig 2.**
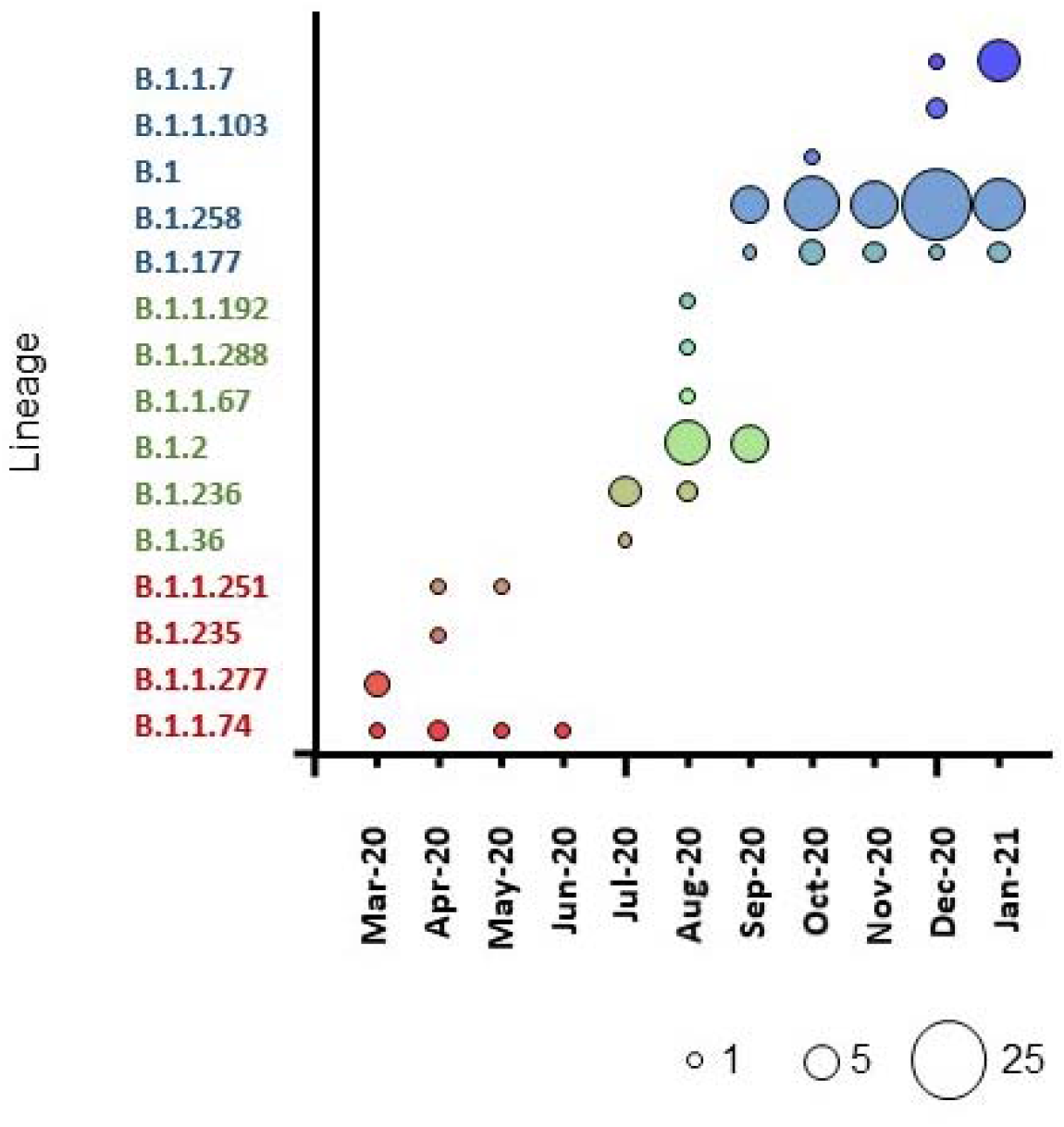
Bubble chart illustrating the pattern of SARS-CoV2 lineages identified per month. The size of the circle is proportional to the number of samples.

**Fig 3.**
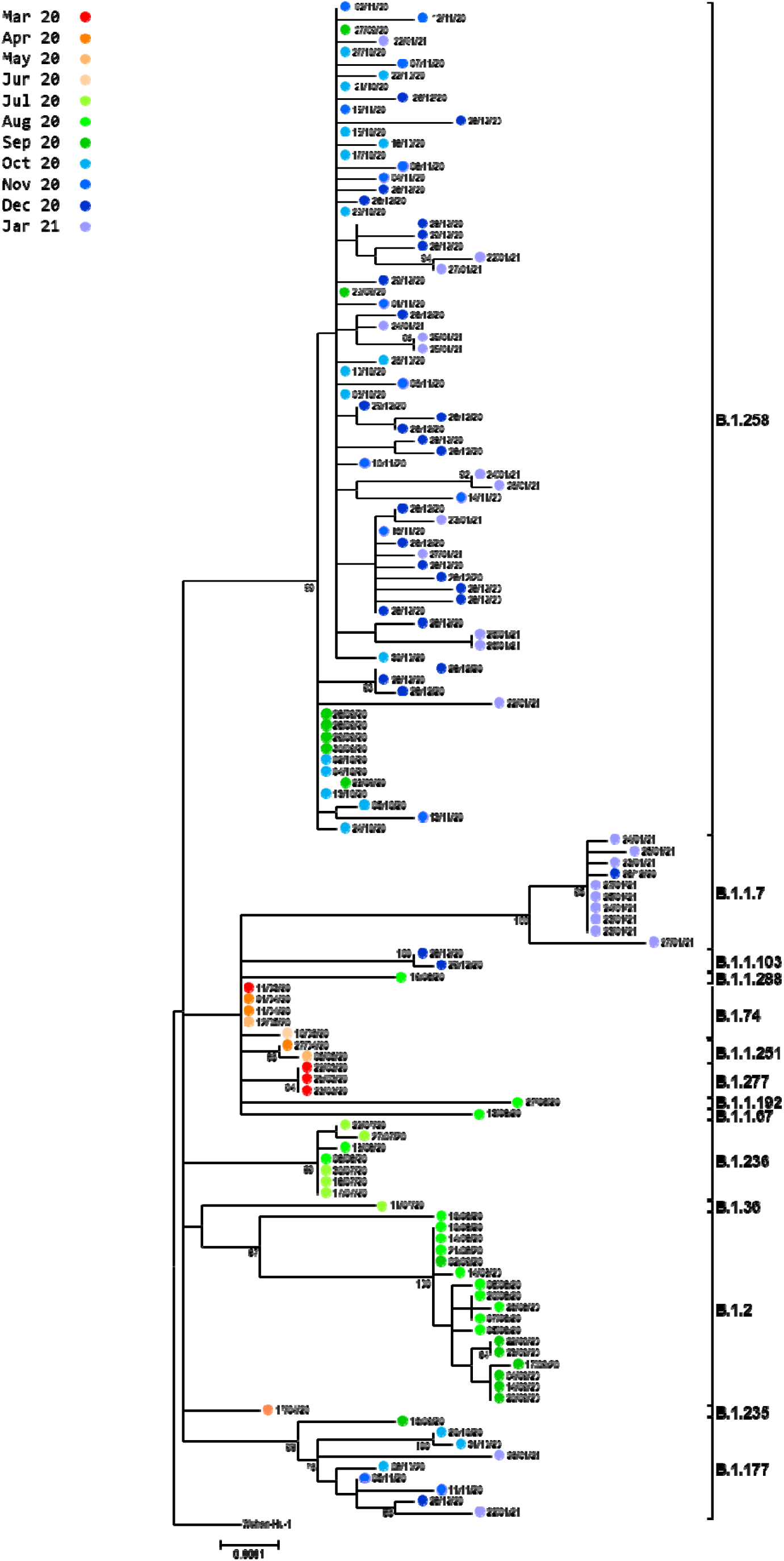
Phylogenetic analysis of full genome sequences of 133 Cypriot strains aligned against the reference genome hCoV-19/Wuhan/Hu-1/2019 (NC_045512.2). Sample names consist of internal laboratory code followed by the date of sampling. The percentages of replicate trees in which the associated taxa clustered together in the standard bootstrap test (1000 replicates) are shown next to the branches. Only bootstrap values >70% are shown.

During the spring wave four variants (B.1.1.74, B.1.1.277, B.1.235 and B.1.1.251) were identified reflecting several independent importation events followed by local transmission. However, concomitant with the decline in reported positive cases following the country-wide lockdown, by the end of June these SARS-CoV-2 variants were no longer detected and none of these re-emerged during the second or the third wave.

Following the re-opening of the airports, the **s**ummer months of July and August were characterized by multiple distinct introductions of a variety of new lineages (B.1.36, B.1.236, B.1.2, B.1.1.67, B.1.1.288, B.1.1.192), which marked the beginning of a small second wave (Fig 3). The majority of the samples belonged to lineage B.1.2 and B1.236 accounting for 86% of the samples in that wave, however, all these lineages again subsequently vanished. The B.1.2 lineage has been otherwise observed only infrequently in Europe, but has been very common in the United States and in Canada [15].

In September, two new lineages were identified for the first time, which were set to dominate the autumn/winter wave until December, namely the B.1.177 and the B.1.258 variants.

The B.1.177 lineage, which is characterized by the A222V spike mutation, was previously been shown to have emerged in Spain in early summer and subsequently became widespread across Europe as well as Canada, accounting for the majority of sequences by autumn 2020 [16,17]. However, no evidence of increased transmissibility of this variant was reported [18]. In Cyprus, this lineage was first identified on the 19^th^ of September and continued to circulate until the end of January at a relatively low but constant frequency fluctuating between 4% and 15 %.

In September, the B.1.258 lineage was introduced, which spread rapidly and largely dominated the autumn/winter wave with a peak prevalence of 86% during the months of November and December.

All samples belonging to the B.1.258 variant identified in Cyprus contain the six-nucleotide ΔH69/V70 deletion in the S gene, which was recently proposed to be labelled B.1.258Δ [19].

This deletion, which has been shown to enhance viral infectivity [20] has arisen at least six times independently and frequently followed receptor binding AA replacements (i.e. N501Y, N439K, Y453F) ([19]). In addition, the B.1.258 variant is characterised by the N439K mutation that confers an increased binding affinity to the hACE2 receptor and leads simultaneously to immune escape from a panel of neutralizing monoclonal antibodies as well as from sera of persons recovered from infection [21].

By the end of December, the B.1.1.7 UK variant, also known as 20I/501Y.V1 and Variant of Concern 202012/01 (VOC-202012/01), was identified for the first time in the Cypriot sample set. This lineage was first reported on the 20^th^ December in the United Kingdom and drew immediate attention due to its rapid spread and increased transmissibility [22]. The earliest sampled genome of the B.1.1.7 variant dates back to the 20^th^ of September. Since then it has grown rapidly in frequency in the UK becoming the most prevalent lineage there with a similar development also observed in a variety of other European countries, including Ireland, Spain and Greece [23]. It is characterised by an unusual large number of genetic changes including the receptor-binding domain and it is speculated that it may have originated in a chronically infected immunocompromised person. In our sample set the frequency of this variant increased in a similar rapid manner from 3.4% in December to 38 % by the end of January despite declining case numbers highlighting its superior fitness.

## Conclusions

Whole genome sequencing of viral specimens following molecular diagnosis is a powerful analytical tool of molecular epidemiology that can critically assist in resolving chains of transmission, identifying of new variants or assessing pathogen evolution and allows a real-time view into the dynamics of a pandemic [24]. In this study we describe for the first time the changing pattern of circulating SARS-CoV-2 lineages in Cyprus since its appearance between March 2020 and the end of January 2021. Distinct lineages of SARS-CoV-2 contributing to three separate waves of infections reflective of the epidemiological pattern were observed.

The global real-time tracing of the viral spread through whole genome sequencing has led recently to the identification of three major Variants of Concern (VOC). Lineage B.1.1.7 (20I/501Y.V1, VOC-202012/01) better known as the UK variant possesses an unusual high number of mutations is believed to be more transmissible than the wild-type virus. Lineage B.1.351 (501.V2, 20H/501Y.V2, VOC-202012/02) first detected and reported in South Africa in early October 2020, shares several mutations with B.1.1.7 and is feared to reduce to some extend vaccine effectiveness [25]. In Brazil, the P.1 (20J/501Y.V3, VOC-202101/02) lineage emerged in December 2020 [26]. The P.1 lineage contains ten mutations in the spike protein that may affect its ability to be recognized by antibodies [27,28].

Of these three VOC only the B1.1.7 variant has so far been identified in Cyprus, which after being encountered for the first time at the end of December was able to reach 37% prevalence within one month. The majority of the Cypriot samples analysed belonged to the B.1.258Δ lineage, which has been first detected at the beginning of August in Switzerland and has since then spread to numerous European countries where it became one of the most prevalent lineages by December including the Czech Republic, Slovakia and Sweden [19,29]. The N439K receptor binding motif characteristic of this lineage has previously been reported to confer an increased binding affinity to the hACE2 receptor and simultaneously leading to immune escape from a panel of neutralizing monoclonal antibodies as well as from sera of persons recovered from infection [21]. The association of mutations found in the B.1.258Δ lineage with increased fitness and immune evasion along with the high prevalence in several European countries stipulates further characterisation of this variant. A continuous surveillance of SARS-CoV2 by whole genome sequencing continues to be critical for timely detection of emerging variants, identifying transmission modes and guiding public health intervention.

## Data Availability

All sequences obtained for this study were deposited at the GISAID EpiCov database (www.gisaid.org).

